# SARS-CoV-2 Receptor Binding Domain IgG Response to AstraZeneca (AZD1222) COVID-19 Vaccination, Jamaica

**DOI:** 10.1101/2021.10.22.21265401

**Authors:** Ynolde E. Leys, Magdalene Nwokocha, Jerome P. Walker, Tiffany R. Butterfield, Velesha Frater, Tamara K. Thompson, Mark Anderson, Gavin A. Cloherty, Joshua J. Anzinger

## Abstract

The Caribbean region is lacking an assessment of the antibody response and side effects experienced after AstraZeneca COVID-19 vaccination (AZD1222). We examined SARS-CoV-2 spike receptor binding domain (RBD) IgG levels and reported side effects in a Jamaican population after AZD1222 vaccination. Median RBD IgG levels for persons without evidence of previous SARS-CoV-2 infection were 43.1 bIU/mL after 3-7 weeks post first dose, rising to 100.1 bIU/mL 3-7 weeks post second dose, and falling 46.9 bIU/mL 16-22 weeks post second dose. The median RBD IgG level 2-8 weeks after symptom onset for unvaccinated SARS-CoV-2 infected persons of all disease severities was 411.6 bIU/mL. Common AZD1222 side effects after first dose were injection site pain, headache and chills. Most persons reported no side effects after second dose. AZD1222 is widely used across the English-speaking Caribbean and the study provides evidence for its continued safe and effective use in vaccination programs.

## Text

The antibody response to COVID-19 vaccination is considered to be of critical importance for protection from COVID-19, especially severe manifestations.^1^ Generally, antibody responses to vaccination can differ depending on the population examined,^2^ and ideally should be assessed for each population. All Caribbean Community (CARICOM) member states have received AstraZeneca vaccines (AZD1222) through COVAX and/or donations, representing the majority of all vaccine types.^3^ Thus far, there have been no studies examining the antibody response to AZD1222 in the Caribbean, necessitating public health decisions based on data from different populations that may use different intervals between vaccine doses.

Jamaica was the first Caribbean country to receive AstraZeneca COVID-19 vaccines via COVAX, with the first administered on March 10, 2021, exactly one year after the first confirmed case in the country.^4^ In this study we examined the antibody response and side effects experienced after AZD1222 for a group of initial vaccinees at the University of the West Indies and University Hospital of the West Indies, Jamaica, and compared responses to unvaccinated SARS-CoV-2 infected persons. Participants were primarily healthcare workers and faculty but also included other persons associated with both institutions. Antibody responses were assessed for persons receiving AZD1222 first doses from March 10-April 27, 2021. Second dose appointments in Jamaica were made available two calendar months after first vaccination, with full dose AZD1222 (5 × 10^10^ viral particles) offered exclusively for these months. This study was approved by the University of the West Indies Mona Campus Research Ethics Committee (CREC-MN.150 20/21).

Antibody responses to vaccination were determined using an Abbott ARCHITECT *i*2000sr instrument (Abbott Laboratories, Abbott Park, Illinois) for the SARS-CoV-2 IgG II Quant and Abbott ARCHITECT SARS-CoV-2 IgM assays: both identified antibodies specific for the spike protein. The SARS-CoV-2 IgG II Quant assay measured antibodies against the spike receptor binding domain (RBD), the domain responsible for binding to ACE2 receptors and a major target of neutralizing antibodies.^5^ SARS-CoV-2 IgG II Quant assay results were reported in WHO binding international units per milliliter (bIU/mL) by multiplying the arbitrary units per milliliter (AU/mL) value by 0.142.^6^ Past infection with SARS-CoV-2 was determined using the Abbott ARCHITECT SARS-CoV-2 IgG assay that identified nucleocapsid-specific IgG. A lower cutoff (≥0.4 S/CO) than the manufacturer’s instructions was used for the SARS-CoV-2 nucleocapsid-specific IgG assay to increase sensitivity as described previously.^7^ All samples in this study were tested with all three assays. SARS-CoV-2 IgG II Quant and IgM assays were considered positive according to the manufacturer’s instructions (≥50 AU/mL and ≥1.0 S/CO, respectively).

A total of 71 AZD1222 vaccinated persons were assessed, 52.1% being males, with an average age of 49.9 years (Table 1). The average time between doses was 9.0 ± 0.8 weeks. Three weeks after first dose all but one person was positive for SARS-CoV-2 RBD IgG and most persons with longitudinal samples showed increased RBD IgG after the second AZD1222 dose that decreased over time (Figure 1A). In participants without serological evidence of previous SARS-CoV-2 infection, median RBD IgG responses were 43.1 bIU/mL (303.5 AU/mL) after 3-7 weeks post first dose, rising to 100.1 bIU/mL (704.6 AU/mL) 3-7 weeks post second dose, and falling to 46.9 bIU/mL (330.3 AU/mL) 16-22 weeks post second dose (Figure 1B and Table 1). The only person not showing evidence of RBD IgG antibodies (<50 AU/mL) after the second AZD1222 dose was notable for an age >80 years (the only participant age >80 years).

**Table 1.** Participant demographic information and SARS-CoV-2 RBD IgG levels

|  | Vaccinated |  |  |  | Infected,<br>Unvaccinated |
| --- | --- | --- | --- | --- | --- |
|  | 3-7 Weeks<br>After First<br>AZD1222 Dose | 3-7 Weeks<br>After Second<br>AZD1222 Dose | 16-22 Weeks<br>After Second<br>AZD1222 Dose | All Time<br>Points After<br>AZD1222 | 2-8 Weeks After<br>Symptom Onset* |
| *n* | 39 | 21 | 36 | 71 | 21 |
| Male | 66.7% | 76.2% | 66.7% | 52.1% | 52.4% |
| Mean age ± st. dev | 50.4 ± 16.1 | 54.8 ± 14.5 | 51.4 ± 16.6 | 49.1 ± 15.5 | 54.1 ± 17.4 |
| SARS-CoV-2 RBD<br>IgG AU/mL (IQR),<br>All Samples | 305.8<br>(91.8-792.9) | 827.0<br>(310.5-1,978.9) | 342.0<br>(168.2-681.0) | N/A | 2,898.9<br>(611.1-11,141.1) |
| SARS-CoV-2 RBD<br>IgG AU/mL (IQR),<br>Excluding Previous<br>Infection | 303.5<br>(91.8-606.2) | 704.6<br>(310.5-1,978.9) | 330.3<br>(146.2-607.1) | N/A | N/A |

\*2-8 week after positive SARS-CoV-2 PCR test for asymptomatic infections.

**Figure 1.**
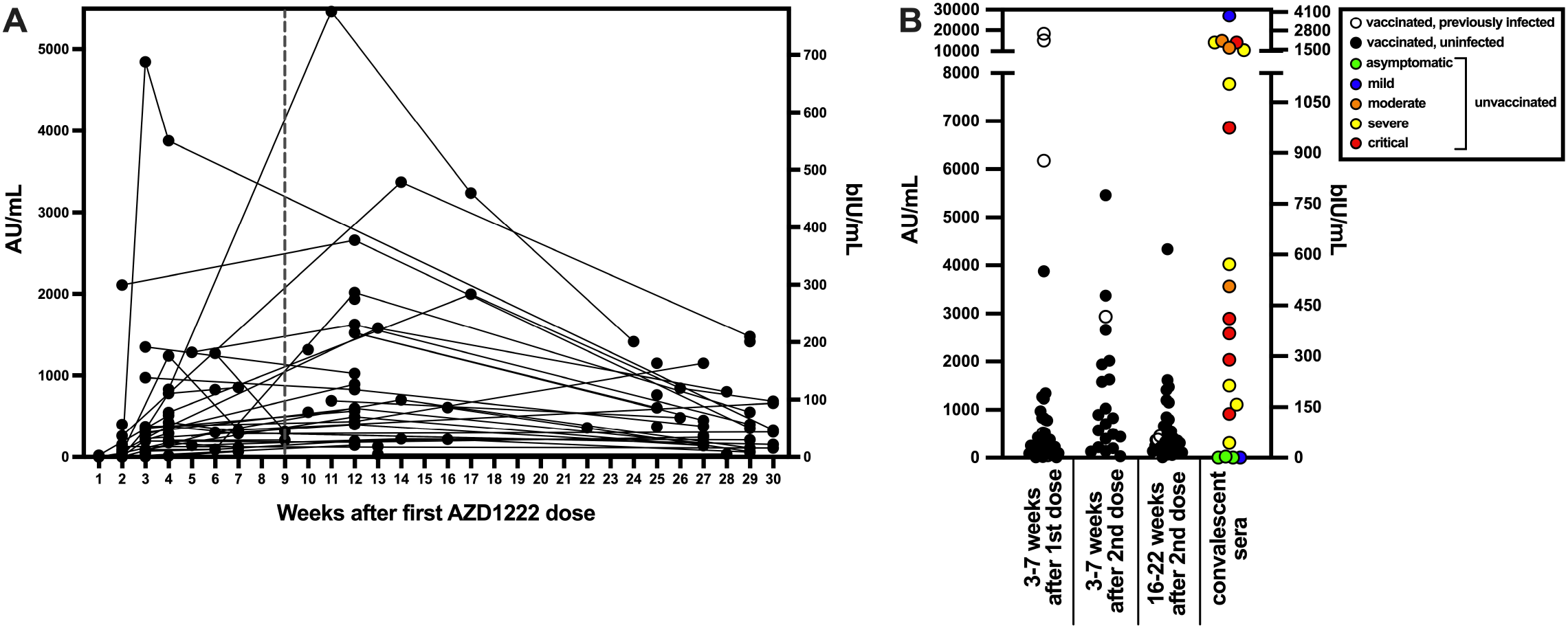
SARS-CoV-2 RBD IgG antibody levels after AZD1222 vaccination and/or SARS-CoV-2 infection. SARS-CoV-2 RBD IgG was measured with the Abbott ARCHITECT SARS-CoV-2 IgG II Quant assay with results reported as arbitrary units per milliliter (AU/mL) on the left y-axis and WHO binding international units per milliliter (bIU/mL) on the right y-axis. A) Sera from all time points collected after AZD1222 vaccination was assessed for SARS-CoV-2 RBD IgG levels. The vertical dotted line indicates the average time of second AZD1222 dose. All data points to the right of the vertical dotted were after second AZD1222 dose B) Sera was collected from AZD1222 vaccinated persons 3-7 weeks after first and second dose, 16-22 weeks after second dose, and for unvaccinated SARS-CoV-2 PCR-confirmed persons 2-8 weeks after symptom onset (or PCR confirmation for asymptomatic persons).

Similar to previous studies with AZD1222 and Pfizer-BioNTech COVID-19 (BNT162b2) vaccines,^8,9^ most vaccinated persons with evidence of previous SARS-CoV-2 infection showed greater RBD IgG levels after first dose than persons vaccinated without evidence of previous SARS-CoV-2 infection (Supplemental Figure 1). The three greatest RBD IgG responses after AZD1222 vaccination were from persons with SARS-CoV-2 PCR-confirmed infection within 2 months prior to receiving the first dose (Supplemental Table 1). In total, twelve persons showed serological evidence of SARS-CoV-2 infection, with four persons experiencing COVID-19 (three SARS-CoV-2 PCR-confirmed and one previously antibody positive) and eight not reporting COVID-19. One person showed serological evidence of SARS-CoV-2 infection after first but before second AZD1222 dose and three persons showed evidence of SARS-CoV-2 breakthrough infection between 3 and 18 weeks after the second dose of AZD1222 (Supplemental Figure 2). None of the persons reported COVID-19 symptoms. Positive SARS-CoV-2 IgM results were generally not observed after AZD1222 vaccination (Supplemental Figure 3) and index values were much lower than we previously reported for SARS-CoV-2 natural infection.^10^

For comparison to AZD1222 vaccination, the RBD IgG response was assessed with convalescent sera from 21 unvaccinated SARS-CoV-2 PCR-confirmed persons, with 52.4% male, and an average age of 54.1 years (Table 1). RBD IgG responses were assessed 2-8 weeks after symptoms onset (or positive PCR test for asymptomatic infections) for unvaccinated persons infected during the initial (April 2020-January 2021) and third waves (July 2021-September 2021) (Table 1 and Figure 1B). Both waves were grouped together, as median RBD IgG responses were not statistically different (Mann-Whitney test, *p* > 0.999) when comparing severe-critical infections for the two waves (only severe-critical infection data was available for the third wave). The median RBD IgG antibody response for unvaccinated PCR-confirmed SARS-CoV-2 infected persons was 411.6 bIU/mL (2,898.9 AU/mL) when including all WHO disease severities. Higher RBD IgG antibody levels were correlated with disease severity as assessed by Spearman’s correlation (ρ = 0.44, *p* = 0.04), in agreement with previous observations.^11^ No correlation was identified for RBD IgG levels and age or sex for unvaccinated SARS-CoV-2 infected persons as assessed by Spearman’s correlation.

Most persons reported side effects and treatment after first AZD1222 dose, whereas after second dose fewer persons reported side effects and they were of shorter duration (Table 2). Spearman’s correlation indicated that myalgia, arthralgia and eye pain were each individually correlated with RBD IgG levels (ρ = 0.385, *p* = 0.02; ρ = 0.374, *p* = 0.03, ρ = 0.368, *p* = 0.03, respectively) after first dose but no correlation between any side effect and RBD IgG levels was identified after second dose. Side effects after first dose were mostly similar to those reported for a phase 1/2 safety and immunogenicity study in a UK population, with the exception of lower percentages reporting fatigue (18% vs 70-71%) and myalgia (16% vs 48-60%) in this study compared to the UK study.^12^

**Table 2.** Side effects and treatments after AZD1222 vaccination

|  | 1<sup>st</sup> Dose | 2<sup>nd</sup> Dose |
| --- | --- | --- |
| Side Effects |  |  |
| None | 19.6% (11/56) | 55.8% (24/43) |
| Injection site pain | 46.4% (26/56) | 20.9% (9/43) |
| Headache | 30.3% (17/56) | 2.3% (1/43) |
| Chills | 25.0% (14/56) | 7.0% (3/43) |
| Fever | 23.2% (13/56) | 2.3% (1/43) |
| Arthralgia | 21.4% (12/56) | 4.7% (2/43) |
| Fatigue | 17.9% (10/56) | 20.9% (9/43) |
| Myalgia | 16.1% (9/56) | - |
| Eye pain | 10.7% (6/56) | - |
| Weakness | 8.9% (5/56) | - |
| Paresthesia | 1.8% (1/56) | 2.3% (1/43) |
| Symptom Duration |  |  |
| 12-24 hr | 46.1% (18/39) | 68.7% (11/16) |
| 36-48 hr | 35.9% (14/39) | 18.7% (3/16) |
| >48 hr | 12.8% (5/39) | 12.5% (2/16) |
| Medication |  |  |
| None | 48.2% (27/56) | 60.0% (24/40) |
| Acetaminophen | 50.0% (28/56) | 37.5% (15/40) |
| Aspirin | - | 2.5% (1/40) |
| Panadeine F | 1.8% (1/56) | - |

This study provides the first assessment of the antibody response to AZD1222 vaccination in the Caribbean region, and provides important information related to common side effects experienced. Most importantly, we show that almost all (70/71) AZD1222 vaccine recipients in this study developed SARS-CoV-2 RBD IgG that remained positive up to 16-22 weeks after second AZD1222 dose.

We observed a lower RBD IgG response to AZD1222 than a previous study in the UK that showed a median SARS-CoV-2 RBD IgG of 435 AU/mL for samples collected >21 days after the first AZD1222 dose using the same Abbott SARS-CoV-2 IgG II Quant assay.^8^ In our study we showed a more modest median response of 303.5 AU/mL 3-7 weeks after the first AZD1222 dose that could possibly be explained by a younger age of the UK population examined and/or genetic or other differences between populations.

The average time to second AZD1222 dose in this study was 9 weeks. The duration between doses can affect the antibody response, with previous studies showing that receipt of the second AZD1222 dose at 12 weeks resulted in greater antibody levels compared to boosting at 8 weeks.^13^ More recent data showed that extended periods between first dose and second dose greater >12 weeks is associated with higher antibody titers.^14^ Although extended times from initial dosing to second dose may be beneficial, consideration must be given for the extent of SARS-CoV-2 circulation within populations that may favor shorter intervals between doses.

This study was limited by the modest sample size and neutralizing antibody testing that was not done. While the SARS-CoV-2 IgG II Quant assay measures IgG antibodies against the spike RBD, a key domain associated with neutralization, the assay does not directly measure neutralizing antibodies. However, RBD IgG levels as measured with the Abbott SARS-CoV-2 IgG II Quant have previously been shown to be correlated with neutralizing antibodies,^15^ and recent studies show that RBD IgG antibody levels are associated with vaccine efficacy.^16,17^ These studies examining correlates of protection indicate the potential utility of measuring RBD IgG antibody levels in the broader population. Future assessment of a larger, nationally representative Jamaican population with assessment of neutralizing antibodies would provide more generalizable information and should be considered by public health officials.

In conclusion, our study shows that AZD1222 vaccination is associated with mild side effects in the Jamaican population and almost always results in RBD IgG that is sustained for at least 22 weeks after vaccination, providing evidence-based support for the continued usage of AZD1222 in the English-speaking Caribbean.

## Data Availability

All data produced in the present study are available upon reasonable request to the authors

## Acknowledgements

As Global Infectious Diseases Scholars, Ynolde Leys and Tiffany Butterfield received mentored research training in the development of this manuscript. This training was supported in part by the University at Buffalo Clinical and Translational Science Institute award UL1TR001412 and the Global Infectious Diseases Research Training Program award D43TW010919. The content is solely the responsibility of the authors and does not necessarily represent the official views of the Clinical and Translational Science Institute or the National Institutes of Health.

## Authors’ addresses

Ynolde E. Leys, Velesha Frater, Joshua J. Anzinger, Department of Microbiology, The University of the West Indies, Mona, Kingston, Jamaica. Magdalene Nwokocha, Department of Microbiology, The University of the West Indies, Mona, Kingston, Jamaica. Mark Anderson, Gavin Cloherty, Infectious Diseases Research, Abbott Diagnostics, Abbott Park, IL, USA

