## Supplemental Table 1 for "SARS-CoV-2 Receptor Binding Domain IgG Response to AstraZeneca (AZD1222) COVID-19 Vaccination, Jamaica"

### Supplementary Materials

Supplemental Table 1
Spike RBD IgG response to first AZD1222 dose in persons with evidence of previous SARS-CoV-2 infection

| Study ID | Days After First AZD1222 Dose | Spike RBD IgG, AU/mL | Nucleocapsid IgG, S/CO | Spike IgM, S/CO |
|---|---|---|---|---|
| 7 | 28 | 511.8 | 1.05 | 0.44 |
| 20[#] | 9 | 11,724.5 | 7.43 | 0.58 |
| 25[#] | 29 | 18,314.3 | 8.11 | 9.64 |
| 29[#] | 35 | 15,178.5 | * | * |
| 41 | 35 | 49.8 | 0.57 | 0.1 |
| 45[@] | 21 | 6,178.1 | 3.33 | 1.44 |

[#]SARS-CoV-2 PCR-confirmed infections. PCR confirmation was 36 days, 26 days and 52 days before first AZD dose for study ID number 20, 25, and 29, respectively. [@]SARS-CoV-2 antibody positive 151 days prior to sampling. [*]Insufficient volume for testing. Study ID 20, 25, 29, and 45 reported COVID-19 symptoms. COVID-19 symptoms were not reported for study ID 7 and 41.

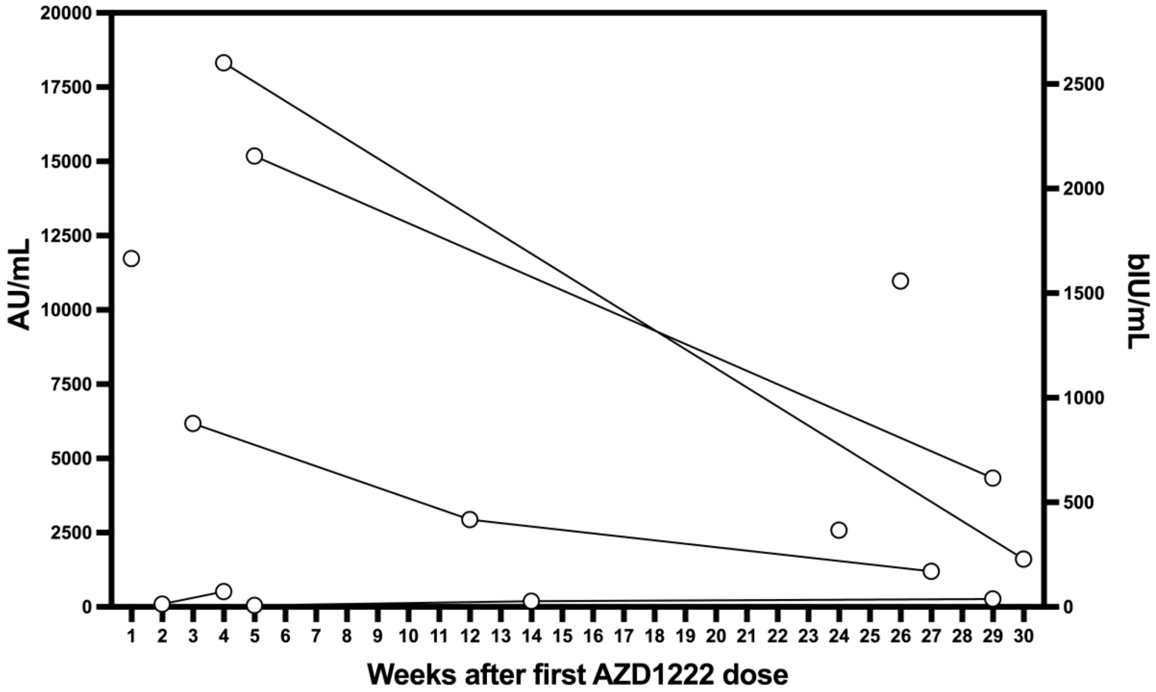


Figure S1. SARS-CoV-2 RBD IgG antibody levels after AZD1222 vaccination for persons with previous evidence of SARS-CoV-2 infection prior to vaccination. SARS-CoV-2 RBD IgG was measured with the Abbott ARCHITECT SARS-CoV-2 IgG II Quant assay with results in arbitrary units per milliliter (AU/mL) on the left y-axis and WHO binding international units per milliliter (bIU/mL) on the right y-axis.

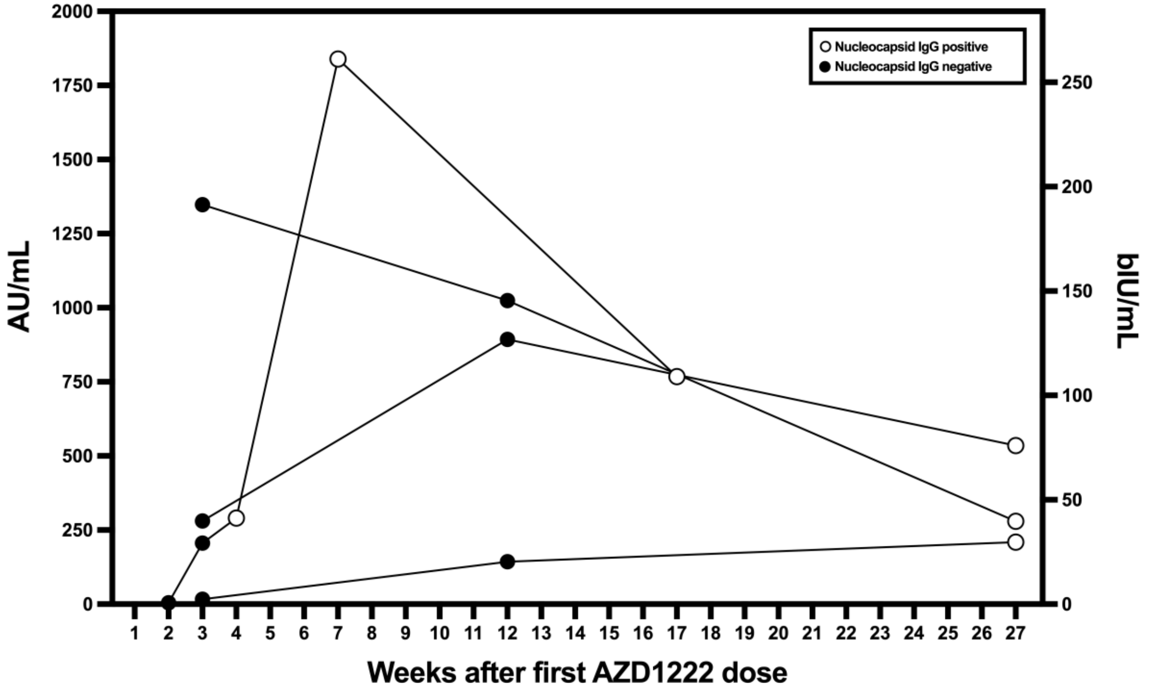


Figure S2. SARS-CoV-2 RBD IgG antibody levels after AZD1222 vaccination for persons with evidence of SARS-CoV-2 infection after vaccination. SARS-CoV-2 RBD IgG was measured with the Abbott ARCHITECT SARS-CoV-2 IgG II Quant assay with results in arbitrary units per milliliter (AU/mL) on the left y-axis and WHO binding international units per milliliter (bIU/mL) on the right y-axis. COVID-19 symptoms were not reported for all persons.

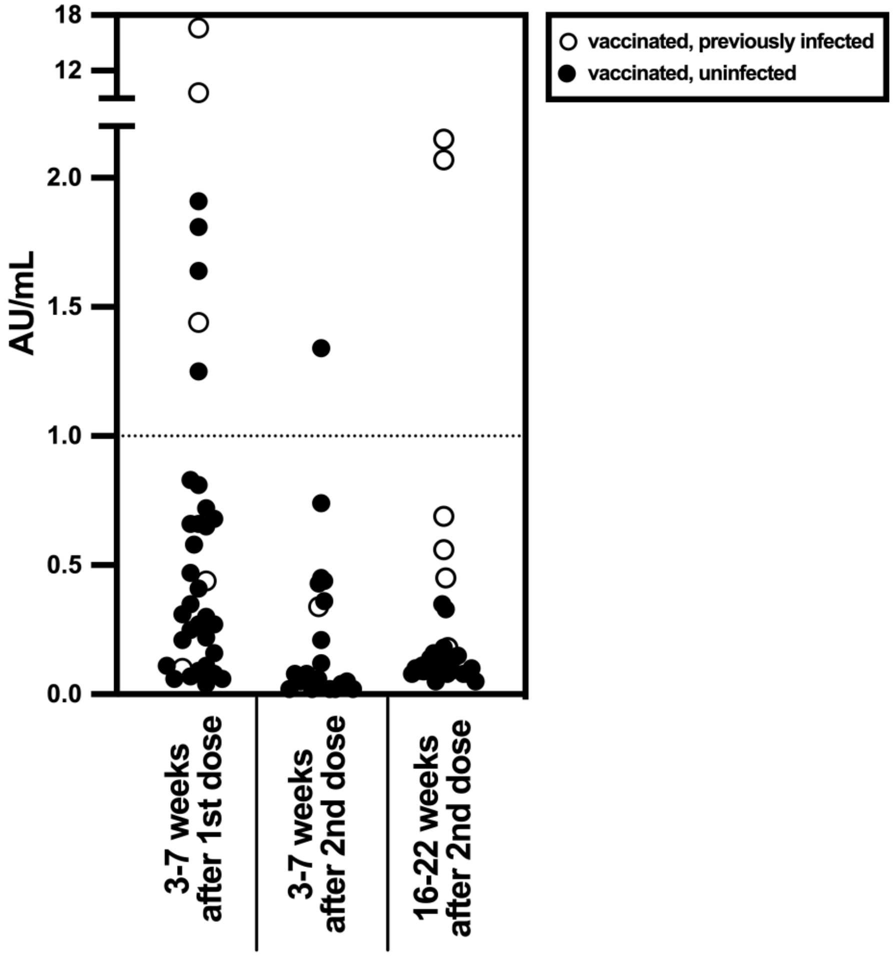


Figure S3. SARS-CoV-2 spike IgM antibody levels after AZD1222 vaccination for persons with and without evidence of previous SARS-CoV-2 infection. SARS-CoV-2 spike IgM was measured with the Abbott ARCHITECT SARS-CoV-2 IgM assay with results in arbitrary units per milliliter (AU/mL). Sera was collected from AZD1222 vaccinated persons 3-7 weeks after first and second dose, and 16-22 weeks after second dose.
